# Multi-contrast machine learning improves schistosomiasis diagnostic performance

**DOI:** 10.1101/2025.01.30.25321353

**Authors:** María Díaz de León Derby, Charles B. Delahunt, Ethan Spencer, Jean T. Coulibaly, Kigbafori D. Silué, Isaac I. Bogoch, Anne-Laure Le Ny, Daniel A. Fletcher

## Abstract

Schistosomiasis currently affects over 250 million people and remains a public health burden despite ongoing global control efforts. Conventional microscopy is a practical tool for diagnosis and screening of *Schistosoma haematobium*, but identification of eggs requires a skilled microscopist. Here we present a machine learning (ML)-based strategy for automated detection of *S. haematobium* that combines two imaging contrasts, brightfield (BF) and darkfield (DF), to improve diagnostic performance. We collected BF and DF images of *S. haematobium* eggs in patient samples from two different field studies in Côte d’Ivoire using a mobile phone-based microscope, the SchistoScope. We then trained separate egg-detection ML models and compared the patient-level performance of BF and DF models alone to Boolean combinations of BF and DF models, using annotations from trained microscopists as the gold standard. We found that models trained on DF images, and almost all BF and DF combinations, performed significantly better than models trained on BF images only. When models were trained on images from the first field study, patient-level classification performance for images from the second study met the WHO Diagnostic Target Product Profile (TPP) sensitivity and specificity for the monitoring and evaluation use case. When we used images from both field studies for the training set, performance of the models was improved. This work shows that multi-contrast imaging can increase information available for classification tasks, while retaining the portability, power, and time-to-results of the TPP’s desired diagnostic. The imaging contrasts used here require no additional sample preparation and do not increase the complexity of the imaging system, and we used off-the-shelf ML models to simplify software engineering. Multi-contrast machine learning offers a practical means to improve performance of automated diagnostics for *S. haematobium*, one that could be applied to other microscopy-based diagnostics.

**Author summary:** Schistosomiasis is a neglected tropical disease that impacts hundreds of millions of people worldwide. Patients with *Schistosoma haematobium* shed parasite eggs in their urine, which can be used as a diagnostic marker of disease. However, identification of those eggs in patient samples normally requires a microscope and trained microscopist. In this work, we show that machine learning models trained on two imaging contrasts, brightfield and darkfield, can improve performance of automated schistosomiasis diagnosis. Using a mobile phone-based microscope (the SchistoScope), we captured brightfield and darkfield images of patient samples during two visits to Côte d’Ivoire and then trained models to detect eggs in the brightfield and darkfield images. When training on images from one visit and testing on images from the other visit, we find that combining the brightfield and darkfield model outputs improved the diagnostic sensitivity and specificity compared to brightfield alone, meeting the target metrics for monitoring and evaluation of schistosomiasis control programs outlined by the World Health Organization. This use of multi-contrast machine learning with a mobile microscope has the potential to improve diagnostic testing for schistosomiasis and could be extended to other neglected tropical diseases.

## Introduction

Schistosomiasis is a neglected tropical disease (NTD) caused by parasitic flatworms that affects more than 250 million people worldwide, with an estimated 800 million people at risk of contracting the disease [1–5]. *Schistosoma haematobium* is one of the main *Schistosoma* species responsible for the disease’s morbidity and mortality. The lack of rapid, portable, and accurate diagnostic tools hinders infection control and elimination efforts in endemic regions.

The standard diagnostic strategy for *S. haematobium* is detection of parasite eggs in urine samples. This method typically involves urine filtration or centrifugation, followed by examination of the sample by a trained expert using light microscopy. These methods are time-consuming and require infrastructure and personnel that are often not available in resource-limited endemic regions. The World Health Organization (WHO) has identified the need for novel diagnostic tools that enable monitoring and evaluation of Schistosomiasis control programs through their Diagnostic Target Product Profiles (TPP) [6]. Ideally, these tools should be portable, use battery-powered equipment, and require minimal training for field workers. Additionally, the amount of time for sample collection, processing, and data interpretation should be less than a working day.

One strategy to facilitate diagnosis of *S. haematobium* and other helminths at the point-of-care is to use portable platforms to image and automatically analyze patient samples. Several groups have developed novel imaging systems that, in combination with machine learning (ML) for image analysis, can be used to identify parasite eggs from urine and stool samples acquired in field settings [7–14]. These devices typically use standard objective lenses and require the use of a computer and mains power.

Additionally, imaging and sample processing times can be long (25-90 minutes) [10, 12] and typically require the samples to be transported to local laboratories foranalysis [9, 11]. ML algorithms for patient-level diagnosis with high-resolution imaging have shown a range of success from 83-96.3% sensitivity and 77-99% specificity, approaching or exceeding the WHO target performance [8, 9, 11, 15].

While existing devices are useful and in some cases meet the WHO TPP sensitivity and specificity criteria, some may be difficult to use for point-of-care detection of schistosomiasis in remote field locations. Their dependence on mains power and bulky hardware are such that they do not meet the requirements for infrastructure, portability and instrumentation outlined in the TPP. Low-cost microscopy with low-resolution imaging is a more workable but under-explored solution for field diagnosis of *S. haematobium*.

In this work, we develop a strategy that uses multi-contrast machine learning based on separate brightfield (BF) and darkfield (DF) images to improve the automated diagnostic performance of the SchistoScope, a mobile phone-based diagnostic platform that can be used to detect *S. haematobium* at the point-of-care [16]. We previously demonstrated that the SchistoScope, a highly portable device (< 1kg) that runs independent of mains power, can be used to simplify *S. haematobium* sample preparation and image acquisition, enabling collection of BF and DF images of patient samples in under 5 minutes. The SchistoScope performs well when compared to conventional on-site light microscopy, as shown in field studies in Ghana and Côte d’Ivoire [16–18], but lack of automated patient-level diagnosis with high sensitivity and specificity has been a limitation.

Here we use images acquired using the SchistoScope with brightfield (BF) and darkfield (DF) illumination to train ML object recognition models using an off-the-shelf YOLOv8 architecture [19] for *S. haematobium* egg detection. Importantly, the ML models we trained can be efficiently deployed on mobile devices. By combining BF and DF imaging contrasts, we boost diagnostic sensitivity and specificity on a patient-level, sufficiently to reach WHO performance metrics for monitoring and evaluation of schistosomiasis control programmes. This strategy of collecting multi-contrast images and using multi-contrast ML to take advantage of the feature information in each contrast may be useful for other portable microscopy systems performing image-based diagnoses.

The use of darkfield imaging for ML-based disease identification and image classification has shown promise in other fields [20–31]. However, to our knowledge, this is the first demonstration of its use as a means to improve diagnostic performance in the context of limited data for diagnosis of neglected diseases.

## Contributions

1. We demonstrate an automated diagnostic strategy that meets WHO requirements for monitoring and evaluation use cases, including sensitivity and specificity, portability, no mains power, and time-to-result.
2. We show that combining ML models trained on brightfield and darkfield images of patient samples taken with a mobile phone-based microscope can improve schistosomiasis diagnostic performance.
3. We collect and annotate a dataset of brightfield and darkfield images of *S. haematobium* that can be used for continued development of machine learning models.

## Materials and methods

### Ethics statement

This work contains patient data from two separate studies conducted in Côte d’Ivoire. The first study was conducted in March 2020 in the Azaguié region of Côte d’Ivoire [17]. Ethical permission for this study was granted by the Centre Suisse de Recherches Scientifiques en Côte d’Ivoire, Abidjan, Côte d’Ivoire (#054-19) and the University Health Network, Toronto, Canada (REB #14-8128). Permission was granted by the local Health District officer. School-age children between 5 and 14 years were invited to participate, and both signed parental consent and the children’s assent were required for inclusion.

The second study was conducted in November 2021, in the Koubi village near the Tiébissou district in Côte d’Ivoire [18]. Ethical permission for this study was granted by the local Health District officer, from the Comité National d’Éthique des Sciences de la Vie et de la Santé, Abidjan, Côte d’Ivoire (REB #186-21) and the University Health Network, Toronto, Canada (REB #21-5582). Community members over 5 years old were asked to participate. Adults provided written consent, and children were included if they assented and had written consent from a parent or guardian.

### Sample processing and image acquisition

Sample processing and image acquisition with the SchistoScope are illustrated in Fig 1A and described in more detail in [16]. For each patient, urine samples were collected in plastic cups and loaded into a 10mL syringe (Fig 1A.i.). The syringe was connected to a custom injection-molded plastic capillary designed to trap *S. haematobium* eggs within an imaging window. The capillary has a rectangular cross-section that tapers down from a height of 200μm at the inlet to 20μm near the outlet hole, trapping and concentrating eggs and other debris as the urine flows through the capillary and exits through the outlet (Fig 1A.ii.). After filtration, the capillary is inserted into the SchistoScope and image acquisition begins. For this, the capillaries are translated in one axis, and images of six distinct fields of view (FOV) are acquired. Each FOV is imaged using both BF and DF illumination.

**Fig 1.**
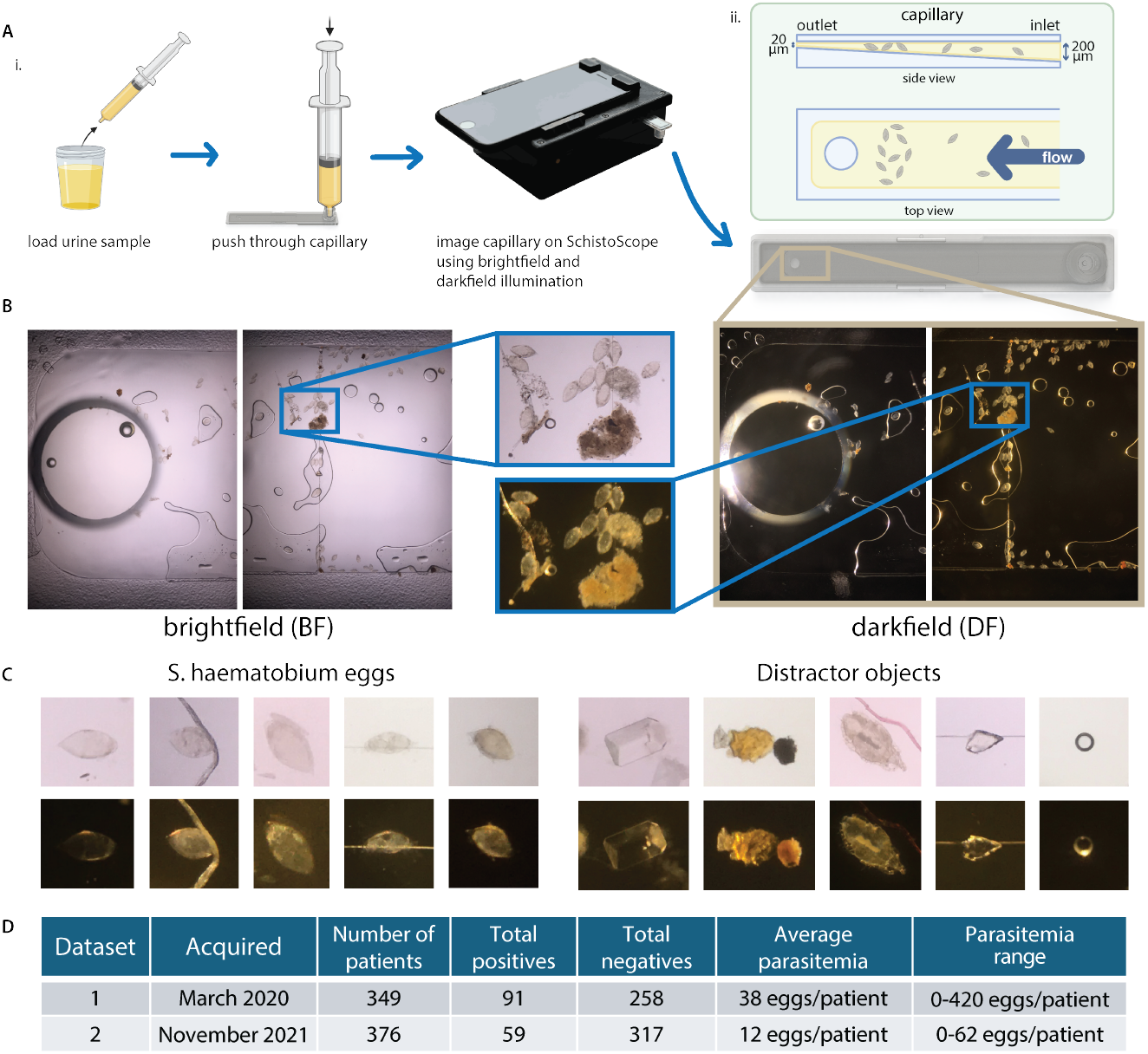
Sample processing, image acquisition and dataset information. **A**: Diagram showing urine sample processing using a capillary and image acquisition with SchistoScope (A.i.). Diagram showing capillary dimensions and egg trapping (A.ii.). Partially created with BioRender.com **B**: Example images in BF and DF of two fields of view of a capillary containing *S. haematobium eggs* and other debris. **C**: Examples of *S. haematobium* eggs and distractor objects trapped in capillaries and imaged with the SchistoScope. **D**: Information about Datasets 1 and 2, which were collected in March 2020 and November 2021, respectively, in different field sites in Côte d’Ivoire, with total number of patients, positive and negative patient count, average parasitemia and parasitemia range shown here.

We implemented BF imaging with an LED illuminator below the sample and DF imaging with an LED illuminator above the sample, oriented at an angle such that unscattered light is not collected by the camera lens. To capture BF and DF images of a single FOV, we turn on the BF illuminator, capture an image, turn off the BF illuminator, turn on the DF illuminator, capture an image, and then turn off the DF illuminator. The entire imaging sequence, including autofocus, takes an average of 60 seconds (range 47-72 sec).

The SchistoScope images are 4032 x 3024 pixels, with pixel pitch ≈ 1 μm/pixel. The optical resolution of the SchistoScope is estimated to be <5 μm [16]. Example images of two distinct FOV for one capillary, captured in BF and DF, are shown in Fig 1B. Due to the tapered design of the capillaries, most eggs are trapped in the region near the outlet hole, corresponding to the first two imaged FOV in each capillary. Examples of *S. haematobium* eggs and “distractor objects”, non-egg debris in urine samples, that were trapped in capillaries and imaged are found in Fig 1C.

### Dataset preparation

The images used for this work were acquired during two separate visits to the *S. haematobium*-endemic regions in Côte d’Ivoire. We created two datasets using the data from these two visits, described below. These datasets are fully described in [32], and will be made publicly available.

The first field visit was completed in March of 2020, in the Azaguié region in Côte d’Ivoire, as described in [17]. Of the 345 individuals tested at this site, 91 patients were positive for *S. haematobium* via light microscopy, most with light-to moderate-burden infections. Only three samples contained more than 50 eggs per 10 mL of urine, meeting the WHO criteria for a high-burden infection [33]. The average parasitemia (number of eggs per positive patient) in this dataset was 38 eggs/patient, as shown in Fig1D. The parasitemia range was 0-420 eggs/patient. Since most eggs are found in the two FOVs closest to the capillary outlet hole for each patient sample, we included those images in the first dataset, regardless of whether or not they contained eggs. We then included, for training only, any additional FOVs that contained eggs (which only happened in samples with very high parasitemia). This resulted in a dataset of 748 total images for each contrast (BF and DF). Of those images, 186 BF images and 188 DF images contained annotated *S. haematobium* eggs. We will refer to the images from this field visit as “Dataset 1”.

The second field visit occurred in the Koubi village near the Tiébissou district in Côte d’Ivoire in November of 2021, as described in [18]. Of the 376 individuals tested at this site, 59 patients were positive for *S. haematobium* via light microscopy, most with low-burden infections. The average parasitemia is 12 eggs/patient and the parasitemia range is 0-62 eggs/patient, both lower than those of Dataset 1. We included the images from the first two FOV for all patients, resulting in 752 images per illumination contrast. Of those images, 99 BF images and 98 DF images contained annotated *S. haematobium* eggs. We refer to the images from this field visit as “Dataset 2”.

### Image annotation

Patient sample images were annotated for the presence of *S. haematobium* eggs by a microscopist with experience in egg identification. These annotations were then verified by another microscopist. In cases of disagreement, a third microscopist was consulted. To carry out the annotations, each image was opened in Microsoft paint, and the center of each visible egg was labelled with a blue dot. Objects that the annotator was unsure of and needed consultation with the second annotator were marked with a red dot. Unlabelled objects in the images are considered distractor objects, some of which are shown in Fig1C.

### ML model training

Due to the relatively small size of our dataset, we used transfer learning to fine-tune pre-trained models to detect Schistosoma eggs (Fig 2A). We chose YOLOv8, developed by Ultralytics and pre-trained on the COCO 2017 dataset, in part because it can be exported to formats such as ONNX and TensorFlow Lite for use on mobile devices [19]. To fit the YOLOv8 input image size of 640×640 pixels, we cropped our 4032×3042 pixel images into 30 individual, partially overlapping, image tiles. We trained the YOLOv8 model using the “detect” task and the following training parameters: stochastic gradient descent optimizer, learning rate of 0.01, and batch size of 16.

**Fig 2.**
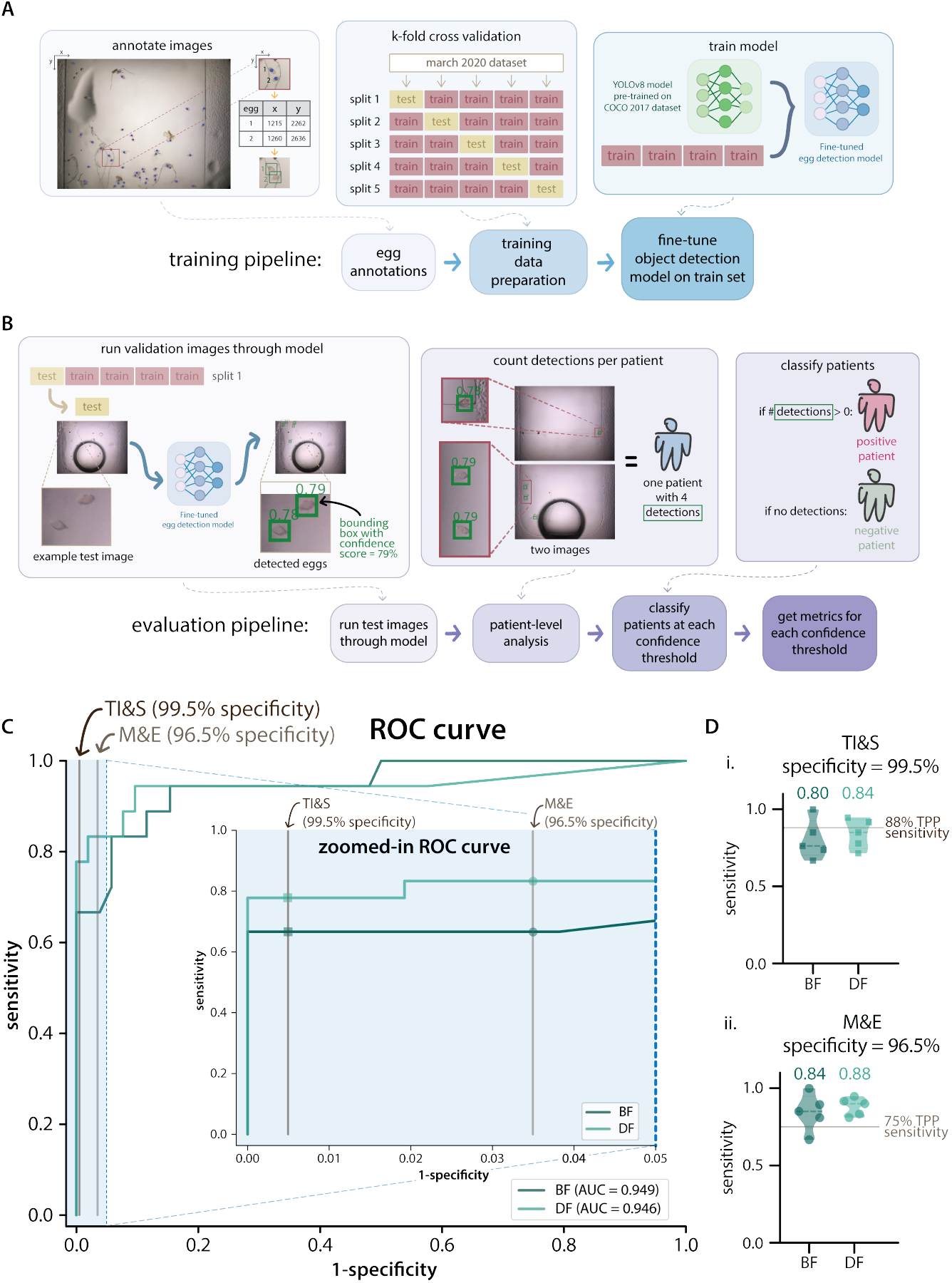
Dataset preparation, ML model training and evaluation of Dataset 1 k-fold splits. **A**: Diagram of the ML model training pipeline. First, *S. haematobium* eggs are annotated in dataset images. For Dataset 1, the patients are then split into 5 folds containing different subsets of training and test data. Transfer learning is done by fine-tuning the ML models (YOLOv8 pre-trained on the COCO 2017 dataset) using the training set for each split. **B**: Diagram of the model evaluation pipeline. After training, the test images are run through the trained model, generating bounding boxes surrounding detections with a confidence score assigned by the model. The number of detections above a certain confidence score threshold are counted for each patient. Each patient is represented by the first two images of a capillary. Subsequently, patients are classified as positive or negative depending on the presence or absence of detections with a confidence score above a given threshold. Sensitivity and specificity metrics are calculated on a patient population level. **C**: Full and zoomed-in receiver operator characteristic (ROC) curves for the first split of the data of Dataset 1, showing results for the BF and DF ML models and the area under each curve. The partial ROC curve is displayed as an inset of the full curve, it shows specificity values from 95% to 100%. The vertical lines indicate the targeted specificity for the transmission interruption and surveillance (TI&S) and monitoring and evaluation (M&E) TPP use cases (99.5% and 96.5%, respectively). **D**: Violin plots showing the patient-level sensitivity values for the 5 splits of Dataset 1 for the TI&S (D.i.) and M&E (D.ii.) use cases. The mean sensitivity is displayed above each violin and the targeted sensitivity for each use case is shown as a vertical line. D.i. shows the sensitivity at a threshold that resulted in 99.5% specificity. D.ii. shows the sensitivity at a threshold that resulted in 96.5% specificity. BF is brightfield and DF is darkfield.

Having data from two different field studies allows us to use one as a holdout set to evaluate the performance of our trained ML models when tested on unseen data. In this work, we set aside Dataset 2 and used Dataset 1 to explore ML model architectures and different ways to combine BF and DF images, as well as for ML hyperparameter tuning. We eventually used all of Dataset 1 to train a final pair of ML models, one for BF and one for DF images. We then used Dataset 2 as our holdout set, using the data to evaluate the models trained on Dataset 1.

We used 5-fold cross-validation (a standard technique to assess model stability), stratified by patient, during our exploratory model training phase using Dataset 1. We divided the 748 dataset images into five different “splits”, each containing a partially overlapping set of images for training, but a completely different set of images for testing. For each of these splits, we trained ML models on the train set images and then evaluated these models on the test set images, as illustrated in Fig 2A. To ensure that images from the same patient were not split between the train and test sets, images that originated from the same patient sample were assigned to the same “group” during k-fold cross-validation.

To ensure an even distribution of images across the splits with different numbers of eggs and distractor objects, we divided the patients into 8 classes: classes 1-3 were positive patients with images that contained eggs in increasing amounts, classes 4-8 were negative patients that contained distractor objects in increasing amounts. We then used the ‘StratifiedGroupKFold’ function from the scikit-learn Python library [34], which splits the data into folds and assigns to each fold roughly equal proportions of each class and also stratifies by patient (i.e. all of a patient’s images are assigned to one fold).

When training the 5-fold split models using Dataset 1, we trained for 200 epochs. When training the final models using all of Dataset 1 to test on Dataset 2, we trained for 300 epochs. In all training instances, we trained separate object-detection models for the BF and DF images.

### Patient classification

Our trained egg-detection models produced a series of detections in each test image that the model identifies as schisto eggs, with an associated confidence score that goes from 1-100%. These detections are indicated by bounding boxes (Fig 2B). Since patient-level, not object-level, performance is what matters clinically, we converted the object-level detections to patient-level diagnostic classification as follows:

First, after running each individual image (composed of 30 image tiles) through the trained model, we combined the detections from the two images corresponding to each patient (Fig 2B). We then evaluated whether each patient would have been classified as positive or negative as we varied a threshold on the confidence score. A patient was considered positive if there was at least one detected object with a confidence score greater or equal to the threshold in any of the images for a patient. Otherwise, the patient was negative. This method applies the patient diagnosis framework in [35], where the noise floor is set to 0 due to the high accuracy of the detection algorithms used.

The object-level precision-recall curves for all splits of the BF and DF models trained and tested on Dataset 1 are shown in Supplementary Figure 1.

### Evaluation metrics

We evaluated our ML models at the patient-level in the test dataset by calculating sensitivity and specificity. We then compared the results to the target metrics for each schistosomiasis diagnostic use case in the WHO Diagnostic Target Product Profiles (TPP) for schistosomiasis control programmes [6].

The following equations define *sensitivity* (Eq. 1) and *specificity* (Eq. 2):

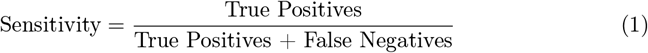

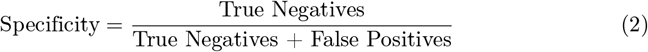

Consistent with convention, True Positive patients are those that were annotated as having *S. haematobium* eggs and were classified as positive by the ML model, while False Negatives are patients that were annotated as having eggs but were classified as negative by the ML model. True Negative patients were both annotated and classified as negative, and False Positives were annotated as negative by human annotators but classified as positive by the ML model.

To show how patient-level sensitivity and specificity depend on the threshold confidence score for model detections, we generated receiver operator characteristic (ROC) curves for each model (BF and DF), which plot sensitivity, or True Positive Rate (TPR), vs 1-specificity, or False Positive Rate (FPR).

To assess the performance of our ML models in the context of Schistosomiasis diagnostics, we evaluated whether we would meet the target metrics established in the WHO TPP [6]. The TPPs are used to guide the development of new diagnostic tools for schistosomiasis for two use cases: (i) Monitoring and Evaluation (M&E) and (ii) Transmission Interruption and Surveillance (TI&S). The TPPs outline the target characteristics of a suitable diagnostic test in categories such as portability, training requirements, throughput, time to results, and clinical sensitivity and specificity. The target sensitivity and specificity for M&E are 75% and 96.5%, respectively. The target sensitivity and specificity for TI&S are 88% and 99.5%, respectively (also shown in Table 1).

**Table 1.**
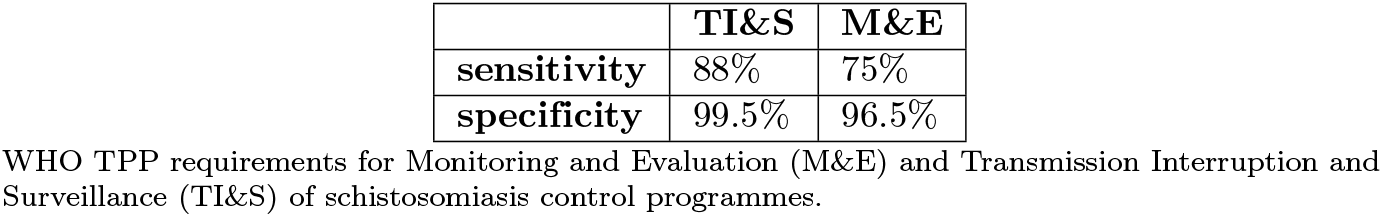
Diagnostic Target Product Profile (TPP) requirements.

We focused our performance analysis on the relevant regions of the ROC curve where specificity was above what is targeted by each WHO use case. Fig 2C shows the full ROC curves for one of the splits (split 1) of the BF and DF ML models trained and evaluated on subsets of Dataset 1, together with a zoomed-in portion of the ROC curve showing the specificity values above 95%. The two vertical lines indicate the specificity values targeted by both of the TPP use cases (96.5% for M&E and 99.5% for TI&S).

To directly compare performance with the TPP use cases, we took the sensitivity at the confidence threshold that resulted in the patient-level specificity targeted by each use case. That is, we set the operating point by requiring that the model meet the specificity in the TPP, then assessed whether it also met the TPP’s sensitivity [35]. Fig 2D shows the sensitivity values for each of the splits of Dataset 1 when evaluated at the targeted specificity for the TI&S (top) and M&E (bottom) use cases. The targeted sensitivity values for each use case are displayed as a horizontal line.

### Multi-contrast combinations

We explored different approaches to combine BF and DF images and assessed whether they would result in improved sensitivity and specificity. The pre-trained YOLOv8 models that we used for transfer learning use 3-channel images as an input. We thus trained separate 3-channel models for BF and DF images and then combined the model outputs with boolean AND or OR, at either object-level or patient-level, for a total of four combination methods. The workflow for these combinations is illustrated in Fig 3A. Merging BF and DF into a 3-channel image (e.g., BF-R, BF-G, DF-B) did not yield good results.

**Fig 3.**
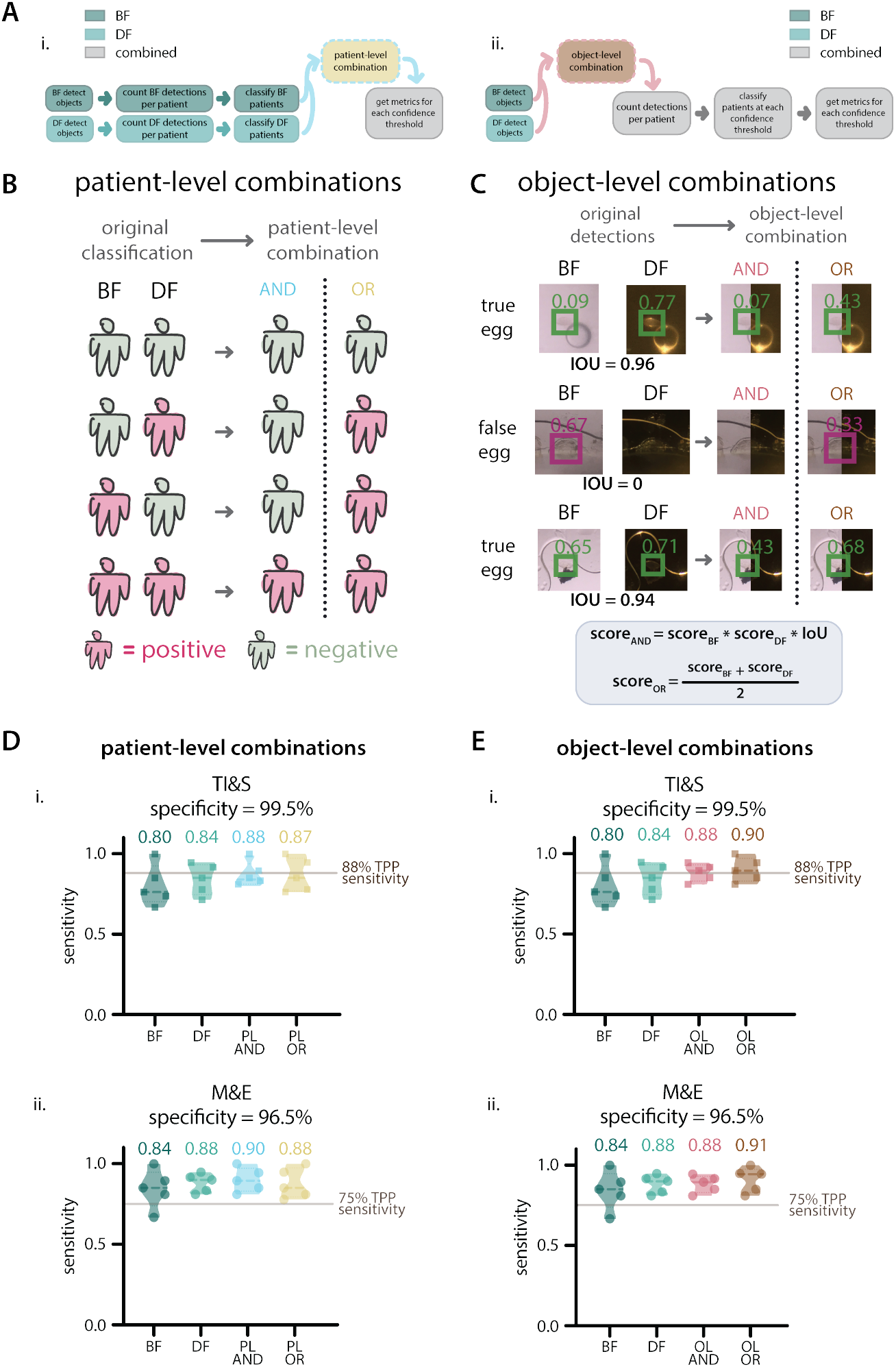
Contrast combination rubrics and patient-level sensitivity on 5-fold splits, Dataset 1. **A**: Combination pipelines. A.i.: Diagram of the patient-level combination pipeline. A.ii.: Diagram of the object-level combination pipeline. **B**: Truth table for patient-level combinations, showing the four possible combinations of patient classifications based on BF and DF models individually, followed by the result after patient-level combinations. Positive patients shown in magenta and negative patients shown in green. **C**: Examples of object-level combinations on three objects in the images, showing original confidence scores assigned by BF and DF models, followed by resulting confidence scores after each combination. Green boxes represent true positive detections and magenta boxes represent false positive detections. **D**: Violin plots showing the sensitivity values after applying patient-level combinations to the 5 splits of Dataset 1 for the TI&S (D.i.) and M&E (D.ii.) use cases. The mean sensitivity is displayed above each violin and the targeted sensitivity for each use case is shown as a vertical line. D.i. shows the sensitivity at a threshold that resulted in 99.5% specificity. D.ii. shows the sensitivity at a threshold that resulted in 96.5% specificity. ‘BF’ is brightfield, ‘DF’ is darkfield, ‘PL AND’ is patient-level AND, ‘PL OR’ is patient-level OR, ‘OL AND’ is object-level AND, ‘OL OR’ is object-level OR. **E**: Violin plots showing the sensitivity values after applying object-level combinations to the 5 splits of Dataset 1 for the TI&S (E.i.) and M&E (E.ii.) use cases. The mean sensitivity is displayed above each violin and the targeted sensitivity for each use case is shown as a vertical line. E.i. sensitivity at a threshold that resulted in 99.5% specificity. E.ii. sensitivity at a threshold that resulted in 96.5% specificity.

For patient-level combinations, we first used the BF and DF model outputs to classify the patients as positive or negative separately for each contrast. After this, we used patient-level AND/OR operations to combine the BF and DF results and arrive at a final diagnosis. For patient-level AND, we called a patient positive only when both BF and DF classified them as positive. For patient-level OR, a positive classification for either BF or DF resulted in a positive combined classification (Fig 3B). After these combinations, we calculated the sensitivity and specificity for the test patient populations and generated ROC curves for the AND and OR cases.

For object-level combinations, we follow the same procedure as above by first separately evaluating images with the BF and DF models, which generates a list of bounding box detections for each contrast. We then apply AND/OR operations at the object level to generate new object scores, as described below, followed by patient-level classification (Fig3A.ii.).

To generate new object scores from the BF and DF detections and scores, we:

i. pair up each individual detection on a BF image with each individual detection on the DF version of that image. Each of these pairs consists of the xy coordinates for the bounding box detection in BF and in DF, as well as their associated confidence scores (score_*BF*_ and score_*DF*_ ).
ii. use the BF and DF xy box coordinates to calculate the intersection over union (IoU) for each detection pair. IoU goes from 0-1 and it measures the overlap between the bounding boxes. If the boxes overlap completely, the IoU is 1. If they are partially overlapping, the IoU is smaller. If the boxes are not overlapping, meaning that a particular object was only detected in one of the contrasts, the IoU is 0.
iii. carry out object-level AND/OR operations to assign a new object score.
  a. For AND, the score is given by:

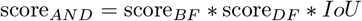 Because the IoU is zero for non-overlapping detections, the object-level AND score eliminates detections that are not represented in both BF and DF. This is a stringent filter, only detections where BF and DF agree on the presence of an egg make it through.
  b. for OR, objects that are only found in BF or DF are not eliminated, but their confidence scores are reduced. To do this, we first eliminate all object pairs that are not overlapping (i.e. pairs with IoU of zero). We then go through the original detection lists for BF and DF, find any detections that are not represented in the combined list, and add them back to the list as “lonely detection pairs”. For these pairs, we assign a confidence score of zero to the missing contrast. For example, if an object is detected only in BF with score=score_*BF*_ , a lonely detection pair is added to the list with score_*BF*_ = score_*BF*_ and score_*DF*_ = 0. After adding the lonely detections, we calculate the object-level OR score as:

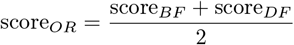 When using the object-level OR combination, we are not removing objects that are only detected in either BF or DF, and by this we hope to avoid eliminating true eggs that were only detected once. However, since we expect a true egg detection to be more likely to be found in both BF and DF, the object-level OR reduces the overall confidence score of lonely detections. Fig 3C shows examples of the resulting scores for object pairs when object-level combinations are applied.

After calculating the object-level AND/OR scores, the patients are classified as positive or negative on a patient-level, based on the presence of combined bounding boxes at a given confidence score threshold. Subsequently, patient-level sensitivity and specificity are calculated and compared to the TPP targets for each use case, as described above.

## Results

### ML model performance on Dataset 1 splits

The BF and DF models were trained on subsets of Dataset 1. We used k-fold splits to better assess their performance before training a set of final models for evaluation using Dataset 2. Results for the BF and DF models are shown in Fig 2, and the results for the BF and DF combinations of those models are shown in Fig 3, all of these results are at the patient-level.

The average sensitivity at the TPP specificity for TI&S for the 5 splits was higher for DF (84%) than for BF (80%). The targeted sensitivity for this TPP use case is 88%, only one split for BF and two splits for DF reached this requirement. However, for the M&E use case, all of the DF splits and most of the BF splits reached the targeted sensitivity of 75%. For this use case, the average sensitivity was also higher for DF (88%) than for BF (84%).

Using DF alone or combinations of BF and DF models resulted a 4-10% increase in mean sensitivity at the targeted TPP specificity values. Notably, when applying both object-level and patient-level combinations, all of the splits of the March 2020 dataset met the TPP requirements for sensitivity and specificity for the M&E use case. Despite not reaching the targeted TPP sensitivity for the TI&S use case on all splits of the data, both object and patient-level combinations increased the average sensitivity, bringing it closer to the WHO targets.

### ML model performance on hold-out Dataset 2

After model training was complete and we confirmed that the performance on Dataset 1 was adequate, we trained models using all of Dataset 1 and tested them on Dataset 2 as a holdout set. This is the scenario that is most realistic and consistent with future diagnostic work in the field, and does not incorporate any information from the test dataset into the training. The patient-level results are shown in Fig 4. A diagram illustrating the data used for training and testing is shown in Fig 4A.

**Fig 4.**
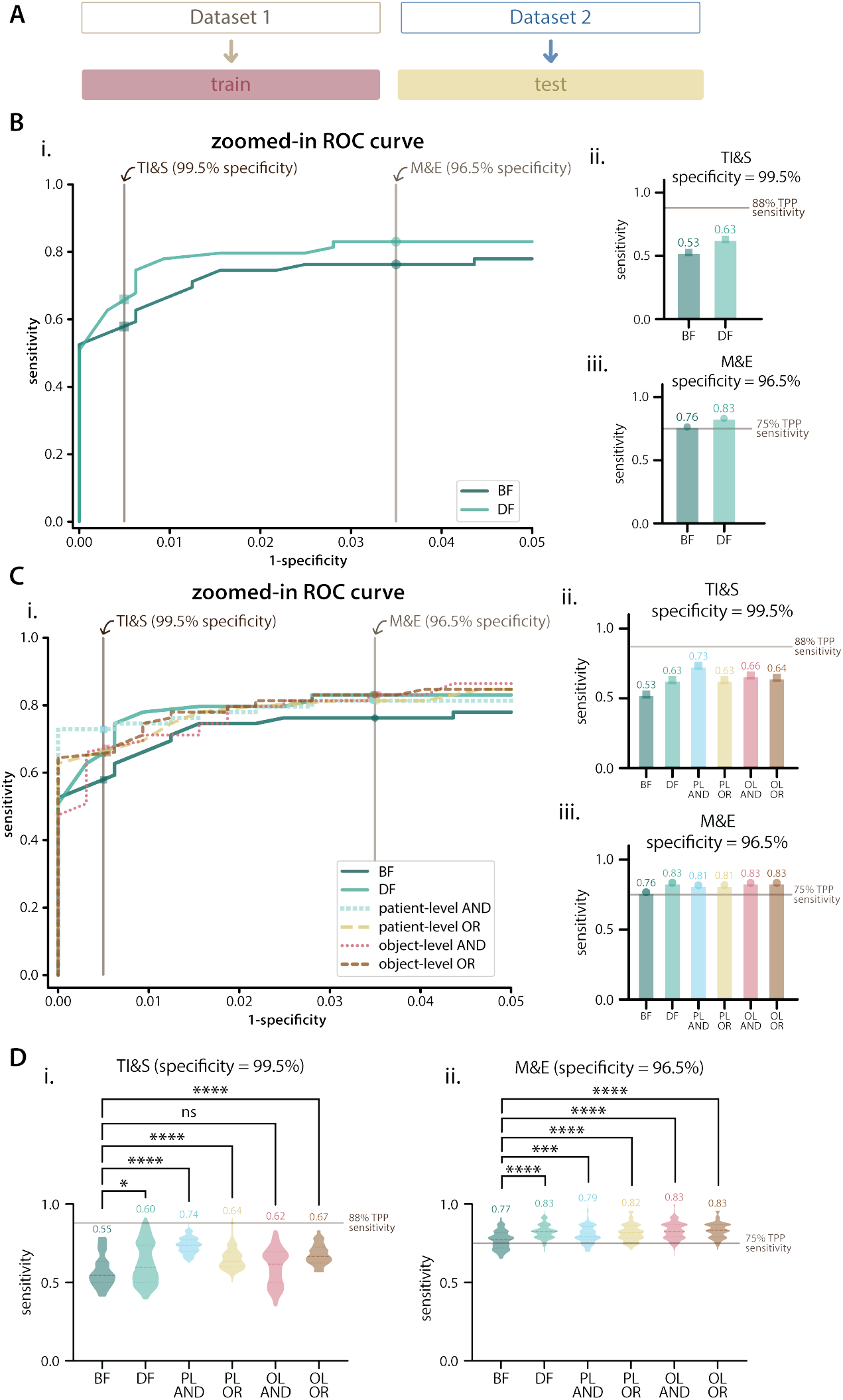
Patient-level results on Dataset 2 as a holdout. **A**: Diagram of data used for training and testing. **B**: Results for brightfield (BF) and darkfield (DF) models trained on Dataset 1 and tested on Dataset 2. B.i. zoomed-in ROC curve showing specificity values from 95% to 100%, with TPP specificity requirements shown as vertical lines. B.ii. and B.iii. patient-level sensitivity for BF and DF models for the TI&S (B.ii.) and M&E (B.iii.) use cases, with sensitivity values for each model displayed above each bar and target sensitivity displayed as a horizontal line. **C**: Results for model combinations on Dataset 2. C.i. zoomed-in ROC curve showing specificity values from 95% to 100%, with TPP specificity requirements shown as vertical lines. C.ii. and C.iii. sensitivity results for BF and DF models and combinations for the TI&S (C.ii.) and M&E (C.iii.) use cases. PL AND is patient-level AND, PL OR is patient-level OR, OL AND is object-level AND, OL OR is object-level OR. **D**: Boostrapping results on the holdout set for TI&S and M&E TPP use cases. The violin plots show the distribution of patient-level sensitivity values at thresholds resulting in the targeted TPP specificity. Bootstrapping was performed for 100 iterations, with sample size = 40% of the patient population. The dashed lines inside violins show the median of the distribution, dotted lines show the quartiles. The median of each distribution is displayed above each violin. We report multiplicity-adjusted p-values, “ns” is p>0.05, ^*^ is p≤0.05, ^***^ is p≤0.001, ^****^ is p≤0.0001.

All models and combinations performed worse when trained on Dataset 1 and tested on Dataset 2, compared to the average performance of k-fold split models trained and tested on subsets of Dataset 1. This is expected, and gives us a better idea of how our trained models would perform with unseen data in future field studies.

Both BF and DF models met the targeted sensitivity for the M&E use case, but they did not meet the targeted sensitivity for the TI&S use case. The DF models performed better than BF when we evaluated the models for both use cases. The combined dataset models resulted in a sensitivity greater or equal to that achieved with the DF models.

The results when the BF and DF models are combined on an object-level and patient-level are shown in Fig 4C. The full ROC curves and AUC of the BF and DF models and combinations is shown in Supplementary Figure 2. None of the contrast combinations achieved the targeted sensitivity for the TI&S use case, but all combinations resulted in a sensitivity greater or equal to that achieved with the BF and DF models separately. The greatest increase was achieved when using a patient-level AND combination.

We used bootstrapping to investigate how our patient-level metrics would have varied had the patient population been a subset of what is in Dataset 2. We iteratively resampled the patient population with replacement, re-running our analysis 100 times on random subsets of 40% of the patients. For each iteration, we calculated the patient-level sensitivity at the threshold resulting in the TPP target specificity, as described above, for the BF-only, DF-only, and combination models. We then used a Kruskal-Wallis test (the non-parametric equivalent of an ANOVA test) with Dunn’s correction for multiple comparisons, to determine whether there were statistically significant differences between the BF and DF models and the combinations. We specifically tested whether the DF model and all the different combinations were significantly different than the BF model (Fig 4D). All statistical analyses were done using GraphPad Prism (version 10.2.2).

The median sensitivity of the bootstrap populations is similar to the sensitivity obtained when testing over the full holdout set. The bootstrapping results show two main things: i. there are statistically significant differences (p ≤ 0.05) between the BF and DF models, as well as between the BF model and most model combinations (with the exception of object-level AND for TI&S). ii. for TI&S, most combination algorithms are more stable than the BF and DF models alone, as seen by their tighter spreads over bootstraps.

### ML model performance on merged datasets (Dataset 3)

Distribution shifts commonly occur when using machine learning for medical tasks, where it is well documented that even small changes from training conditions can lead to changes in performance [35–38]. Given that we saw a decrease in performance when models trained on Dataset 1 were tested on Dataset 2, we wondered if it was due to a distribution shift between the datasets, since they were acquired during different field visits.

As a way to test this, we created a “merged” dataset by combining the images from both datasets into one, which we refer to as “Dataset 3”. If we are affected by distribution shifts, we would expect the models trained and tested a subset of Dataset 3 to perform better than those trained on Dataset 1 and tested on Dataset 2.

We randomly split Dataset 3 into a train set and a test set, as illustrated in Fig 5A.i. By using Dataset 3 we could see how our model would perform in the ostensibly best available case, where maximum training data is used and testing is done in-distribution. The average parasitemia for the test set of Dataset 3 was 26 eggs/patient, and the parasitemia range for the test set was 0-420 eggs/patient, as shown in Fig5B.i. and B.ii.

**Fig 5.**
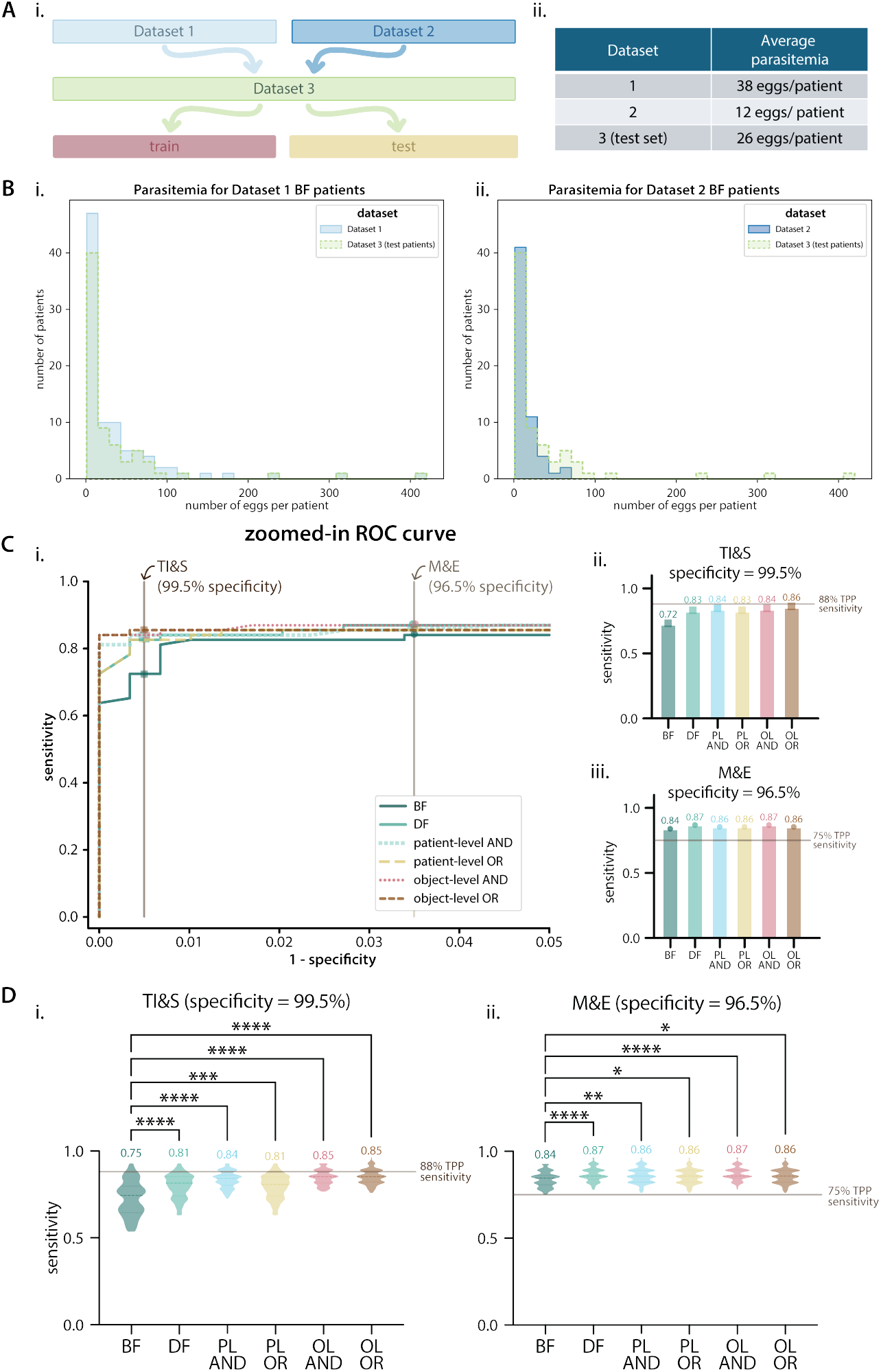
Patient-level results on Dataset 3. **A** Dataset 3 information. A.i.: Diagram of the data used for Dataset 3. A.ii.: Average parasitemia for positive patients on the test set of Dataset 3 compared to the average parasitemia of Datasets 1 and 2. **B**: Histograms showing the distribution of patient-level parasitemia for Dataset 1 (B.i.) and Dataset 2 (B.ii.) compared to the distribution of the test set for Dataset 3 **C**: Patient-level results on the test set of Dataset 3. C.i. zoomed-in ROC curve with specificity values ranging from 95% to 100%, with TPP specificity requirements shown as vertical lines. C.ii. and C.iii. show the patient-level sensitivity results for BF and DF models and combinations for the TI&S and M&E use cases (at thresholds resulting in TPP specificity). The target sensitivity for each use case is shown as a horizontal line. BF is brightfield, DF is darkfield, PL AND is patient-level AND, PL OR is patient-level OR, OL AND is object-level AND, OL OR is object-level OR. **D**: Boostrapping results on the test set of Dataset 3 for TI&S and M&E TPP use cases. The violin plots show the distribution of patient-level sensitivity values at thresholds resulting in the targeted TPP specificity. Bootstrapping was performed for 100 iterations, with sample size = 40% of the patient population. The dashed lines inside violins show the median of the distribution, dotted lines show the quartiles. The median of each distribution is displayed above each violin. We report multiplicity-adjusted p-values, “ns” is p>0.05, ^*^ is p≤0.05, ^***^ is p≤0.001, ^****^ is p≤0.0001.

Overall, the performance of the ML models on Dataset 3 was better than the performance of the models trained on Dataset 1 and tested on Dataset 2. When evaluated at thresholds that met the TPP target specificity, all contrasts and combinations met the TPP target sensitivity for the M&E use case. For the TI&S use case, no contrast or combination met the target TPP sensitivity. The object-level OR combination was the closest, with only 2% lower sensitivity than the target.

We performed bootstrapping to gain insight on the variability of the patient-level metrics. For the TI&S use case, DF and all of the contrast combinations performed significantly better than BF. The object-level combinations (AND and OR) had both the highest median sensitivity and the tightest distributions. Notably, the third quartile for both of these distributions was above the sensitivity targeted by the TPP (88% sensitivity). 29/100 iterations for object-level AND and 30/100 iterations for object-level OR had a sensitivity above the TPP target for TI&S.

Fig 5D.ii. shows the sensitivity distributions for the M&E use case. DF and all of the combinations performed significantly better than BF. Notably, for all of the contrasts and combinations and for all of the 100 iterations, the models had a sensitivity above or equal to that targeted by the TPP (75% sensitivity) at a threshold that resulted in the targeted specificity.

## Discussion

Diagnostic technologies that are simple to use, low cost, and achieve WHO performance metrics are needed to advance schistosomiasis control and elimination goals. The development of mobile phone-based microscopes for image-based diagnosis of *S. haematobium*, such as the SchistoScope, partially achieve those goals through their portability and simplicity. However, the best strategy for automated egg detection and patient diagnosis for mobile microscope with moderate resolution has been unclear, as many existing ML models rely on images collected with high-resolution imaging systems. Moderate resolution systems, including the SchistoScope, may need additional information to achieve the combination of sensitivity and specificity needed for field applications. This paper highlights the potential of DF as a mean to break the zero-sum trade-off between accuracy and practicality, by enabling portable, lower resolution systems, to support high accuracy detection.

We use multi-contrast imaging of patient urine samples containing *S. haematobium* acquired in endemic regions of Côte d’Ivoire using the SchistoScope to train ML models for automated diagnosis. We find that DF models alone and combinations of BF and DF models lead to greater performance than BF alone, which is the typical contrast used to identify eggs with light microscopy. The combinations of BF and DF models meet the WHO target sensitivity and specificity for monitoring and evaluation of schistosomiasis control programmes, with DF consistently showing better performance than BF. A relatively small dataset of less than 1000 images was sufficient to train the models and demonstrate improved diagnostic performance, taking advantage of the availability of pre-trained, off-the-shelf ML models that can be used for fine-tuning to a particular application.

Our multi-contrast machine learning approach benefited from BF and DF images providing complimentary information about the schistosoma eggs, with brightfield contrast reporting light absorption by the sample and darkfield contrast showing scattering by sample edges and other features. Darkfield, or pseudo-darkfield, can be easily (and fairly inexpensively) implemented in a standard light microscope by adding an oblique or annular illumination source, or by blocking illumination angles that are captured by the imaging lens, an example of which is shown in [39]. Hence, DF imaging can be implemented by other groups integrating ML with portable microscopy for diagnosis of *S. haematobium* and other diseases with egg-based diagnostics, such as *Schistosoma mansoni* and soil-transmitted helminths. In fact, DF imaging alone can be helpful for semi-automatic diagnostic strategies where clinicians or field technicians make diagnostic calls based on digitized images of patient samples. Our annotators and clinical collaborators noted that they preferred annotating/evaluating darkfield images because *S. haematobium* eggs are easier to identify in darkfield versus the traditional brightfield contrast.

In cases where a microscopy system supports both BF and DF imaging, these can be combined in relatively simple ways to get better ML results. We showed that simple, boolean combinations of patient-level diagnosis with models trained on images of different contrasts (such as patient-level AND/OR) can lead to improvements in performance. Incorporating additional contrasts, such as differential phase contrast and fluorescence, into portable and low-cost microscopes could provide additional sample information that might further improve multi-contrast machine learning performance. In particular, the autofluorescence of Schistosoma eggs and other parasites makes this an attractive direction for future device development. Improvements in ML model development could also advance the goal of high performance detection with lower resolution images, including altering the model architecture to train on images of both contrasts simultaneously, increasing hyperparameter optimization, and training on more egg images. As the field of ML continues to evolve rapidly, novel models and architectures could also lead to performance improvements, potentially reaching the TI&S target metrics.

An important next step to validate the usefulness of multi-contrast machine learning will be to do live field testing of the ML models loaded onto the SchistoScope or its successor. This will require exporting our ML models to a mobile phone-compatible format to evaluate performance and processing time. Any future field deployment will also require selecting confidence score thresholds in advance and providing patient-level diagnosis based on them. We observed ML model performance improvements when training on the combined dataset (Dataset 3) compared to training on data from one site (Dataset 1) and testing on data from the second site (Dataset 2); this type of variability has also been observed during field testing for other diagnostic products [38]. Given this, future field deployments should consider real-time updates of the ML models to accommodate inter-clinic variability and uneven algorithm accuracy at new sites due to distribution shifts in training and testing populations. It is also worth noting that the WHO TPP for TI&S is in the context of disease elimination, which generally implies lower parasitemia distributions, making it harder to hit the sensitivity targets [35]. We expect our multi-contrast strategy to be even more relevant in this context.

## Conclusion

Mobile phone-based microscopy platforms in conjunction with multi-contrast machine learning and novel sample preparation techniques can be used for rapid, sensitive, and portable diagnosis of *S. haematobium* that meets WHO diagnostic requirements. Performance of ML models to identify Schistosoma eggs can be significantly improved by adding darkfield imaging to standard brightfield microscopes, which requires minimal changes in microscope optics and no additional sample preparation. Multi-contrast machine learning with an additional contrast offers a practical means to improve performance of low-cost, automated diagnostics for *S. haematobium* egg detection and could be applied to other microscopy-based diagnostics.

## Data Availability

The data used in this paper has been made available on the AFRICAI repository in the
Euro-BioImaging Medical Imaging Archive XNAT (https://xnat.health-
ri.nl/app/template/XDATScreen_report_xnat_projectData.vm/search_element/xnat:proje
ctData/search_field/xnat:projectData.ID/search_value/AFRICAI_MICCAI2024_Schistos
omias). The data will also be uploaded and made publicly available through Zenodo.

https://xnat.health-ri.nl/app/template/XDATScreen_report_xnat_projectData.vm/search_element/xnat:projectData/search_field/xnat:projectData.ID/search_value/AFRICAI_MICCAI2024_Schistosomias

## Acknowledgments

We thank the Berkeley Research Computing program at the University of California, Berkeley, for providing the Savio computational cluster resource, which we used to train and evaluate our models.

## Author contributions

- Conceptualization and study design: MDDLD, CBD, DAF
- Field data collection: JTC, KDC, IIB
- Image annotations: ES, MDDLD
- Experiments and data analysis: MDDLD
- ML model training: MDDLD
- Funding acquisition: JTC, IIB, ALLN, DAF
- Supervision: CBD, IIB, ALLN, DAF
- Writing – original draft: MDDLD, CBD, DAF
- Writing – review & editing: all authors

## References

1. McManus DP, Dunne DW, Sacko M, Utzinger J, Vennervald BJ, Zhou XN. Schistosomiasis. Nature Reviews Disease Primers. 2018;4(1):13. doi:10.1038/s41572-018-0013-8.

2. World Health Organization. WHO guideline on control and elimination of human schistosomiasis;. https://www.who.int/publications/i/item/9789240041608.

3. World Health Organization Diagnostics Technical Advisory Group (DTAG). Global report on neglected tropical diseases 2023;. https://www.who.int/publications/i/item/9789240067295.

4. Deol AK, Fleming FM, Calvo-Urbano B, Walker M, Bucumi V, Gnandou I, et al. Schistosomiasis — Assessing Progress toward the 2020 and 2025 Global Goals. New England Journal of Medicine. 2019;381(26):2519–2528. doi:10.1056/nejmoa1812165.

5. Li EY, Gurarie D, Lo NC, Zhu X, King CH. Improving public health control of schistosomiasis with a modified WHO strategy: a model-based comparison study. The Lancet Global Health. 2019;7(10):e1414–e1422. doi:10.1016/s2214-109x(19)30346-8.

6. World Health Organization Diagnostics Technical Advisory Group (DTAG). Diagnostic target product profiles for monitoring, evaluation and surveillance of schistosomiasis control programmes;. https://www.who.int/publications/i/item/9789240031104.

7. Meulah B, Bengtson M, Lieshout LV, Hokke CH, Kreidenweiss A, Diehl JC, et al. A review on innovative optical devices for the diagnosis of human soil-transmitted helminthiasis and schistosomiasis: from research and development to commercialization. Parasitology. 2022;150(2):137–149. doi:10.1017/s0031182022001664.

8. Meulah B, Oyibo P, Bengtson M, Agbana T, Lontchi RAL, Adegnika AA, et al. Performance Evaluation of the Schistoscope 5.0 for (Semi-)automated Digital Detection and Quantification of Schistosoma haematobium Eggs in Urine: A Field-based Study in Nigeria. The American Journal of Tropical Medicine and Hygiene. 2022;107(5):1047–1054. doi:10.4269/ajtmh.22-0276.

9. Meulah B, Oyibo P, Hoekstra PT, Moure PAN, Maloum MN, Laclong-Lontchi RA, et al. Validation of artificial intelligence-based digital microscopy for automated detection of Schistosoma haematobium eggs in urine in Gabon. PLOS Neglected Tropical Diseases. 2024;18(2):e0011967. doi:10.1371/journal.pntd.0011967.

10. Oyibo P, Meulah B, Bengtson M, van Lieshout L, Oyibo W, Diehl JC, et al. Two-stage automated diagnosis framework for urogenital schistosomiasis in microscopy images from low-resource settings. Journal of Medical Imaging. 2023;10(4):044005. doi:10.1117/1.JMI.10.4.044005.

11. Makau-Barasa L, Assefa L, Aderogba M, Bell D, Solomon J, Urude RO, et al. Performance evaluation of the AiDx multi-diagnostic automated microscope for the detection of schistosomiasis in Abuja, Nigeria. Scientific Reports. 2023;13(1). doi:10.1038/s41598-023-42049-6.

12. Ward P, Dahlberg P, Lagatie O, Larsson J, Tynong A, Vlaminck J, et al. Affordable artificial intelligence-based digital pathology for neglected tropical diseases: A proof-of-concept for the detection of soil-transmitted helminths and Schistosoma mansoni eggs in Kato-Katz stool thick smears. PLOS Neglected Tropical Diseases. 2022;16:1–16. doi:10.1371/journal.pntd.0010500.

13. Lundin J, Suutala A, Holmström O, Henriksson S, Valkamo S, Kaingu H, et al. Diagnosis of soil-transmitted helminth infections with digital mobile microscopy and artificial intelligence in a resource-limited setting. PLOS Neglected Tropical Diseases. 2024;18(4):e0012041. doi:10.1371/journal.pntd.0012041.

14. Rubio Maturana C, Dantas de Oliveira A, Zarzuela F, Ruiz E, Sulleiro E, Mediavilla A, et al. Development of an automated artificial intelligence-based system for urogenital schistosomiasis diagnosis using digital image analysis techniques and a robotized microscope. PLOS Neglected Tropical Diseases. 2024;18(11):e0012614. doi:10.1371/journal.pntd.0012614.

15. Oyibo P, Jujjavarapu S, Meulah B, Agbana T, Braakman I, van Diepen A, et al. Schistoscope: An Automated Microscope with Artificial Intelligence for Detection of Schistosoma haematobium Eggs in Resource-Limited Settings. Micromachines. 2022;13(5). doi:10.3390/mi13050643.

16. Armstrong M, Harris AR, D’Ambrosio MV, Coulibaly JT, Essien-Baidoo S, Ephraim RKD, et al. Point-of-Care Sample Preparation and Automated Quantitative Detection of Schistosoma haematobium Using Mobile Phone Microscopy. The American Journal of Tropical Medicine and Hygiene. 2022;106(5):1442 – 1449. doi:10.4269/ajtmh.21-1071.

17. Coulibaly JT, Silue KD, Armstrong M, de León Derby MD, D’Ambrosio MV, Fletcher DA, et al. High Sensitivity of Mobile Phone Microscopy Screening for Schistosoma haematobium in Azaguié, Côte d’Ivoire. The American Journal of Tropical Medicine and Hygiene. 2023;108(1):41 – 43. doi:10.4269/ajtmh.22-0527.

18. Coulibaly JT, Silue KD, Díaz de León Derby M, Fletcher DA, Fisher KN, Andrews JR, et al. Rapid and Comprehensive Screening for Urogenital and Gastrointestinal Schistosomiasis with Handheld Digital Microscopy Combined with Circulating Cathodic Antigen Testing. The American Journal of Tropical Medicine and Hygiene. 2024; doi:10.4269/ajtmh.24-0043.

19. Jocher G, Chaurasia A, Qiu J. Ultralytics YOLOv8; 2023. Available from: https://github.com/ultralytics/ultralytics.

20. Fan L, Huang T, Lou D, Peng Z, He Y, Zhang X, et al. Artificial Intelligence-Aided Multiple Tumor Detection Method Based on Immunohistochemistry-Enhanced Dark-Field Imaging. Analytical Chemistry. 2021;94(2):1037–1045. doi:10.1021/acs.analchem.1c04000.

21. Lippeveld M, Knill C, Ladlow E, Fuller A, Michaelis LJ, Saeys Y, et al. Classification of Human White Blood Cells Using Machine Learning for Stain-Free Imaging Flow Cytometry. Cytometry Part A. 2019;97(3):308–319. doi:10.1002/cyto.a.23920.

22. Hennig H, Rees P, Blasi T, Kamentsky L, Hung J, Dao D, et al. An open-source solution for advanced imaging flow cytometry data analysis using machine learning. Methods. 2017;112:201–210. doi:10.1016/j.ymeth.2016.08.018.

23. Blasi T, Hennig H, Summers HD, Theis FJ, Cerveira J, Patterson JO, et al. Label-free cell cycle analysis for high-throughput imaging flow cytometry. Nature Communications. 2016;7(1). doi:10.1038/ncomms10256.

24. Eulenberg P, Köhler N, Blasi T, Filby A, Carpenter AE, Rees P, et al. Reconstructing cell cycle and disease progression using deep learning. Nature Communications. 2017;8(1). doi:10.1038/s41467-017-00623-3.

25. Nassar M, Doan M, Filby A, Wolkenhauer O, Fogg DK, Piasecka J, et al. Label-Free Identification of White Blood Cells Using Machine Learning. Cytometry Part A. 2019;95(8):836–842. doi:10.1002/cyto.a.23794.

26. Doan M, Case M, Masic D, Hennig H, McQuin C, Caicedo J, et al. Label-Free Leukemia Monitoring by Computer Vision. Cytometry Part A. 2020;97(4):407–414. doi:10.1002/cyto.a.23987.

27. Yaroshenko A, Hellbach K, Yildirim AO, Conlon TM, Fernandez IE, Bech M, et al. Improved In vivo Assessment of Pulmonary Fibrosis in Mice using X-Ray Dark-Field Radiography. Scientific Reports. 2015;5(1). doi:10.1038/srep17492.

28. Willer K, Fingerle AA, Noichl W, De Marco F, Frank M, Urban T, et al. X-ray dark-field chest imaging for detection and quantification of emphysema in patients with chronic obstructive pulmonary disease: a diagnostic accuracy study. The Lancet Digital Health. 2021;3(11):e733–e744. doi:10.1016/s2589-7500(21)00146-1.

29. Emons J, Fasching PA, Wunderle M, Heindl F, Rieger J, Horn F, et al. Assessment of the additional clinical potential of X-ray dark-field imaging for breast cancer in a preclinical setup. Therapeutic Advances in Medical Oncology. 2020;12:1758835920957932. doi:10.1177/1758835920957932.

30. Frank M, Gassert FT, Urban T, Willer K, Noichl W, Schick R, et al. Dark-field chest X-ray imaging for the assessment of COVID-19-pneumonia. Communications Medicine. 2022;2(1). doi:10.1038/s43856-022-00215-3.

31. Partridge T, Astolfo A, Shankar SS, Vittoria FA, Endrizzi M, Arridge S, et al. Enhanced detection of threat materials by dark-field x-ray imaging combined with deep neural networks. Nature Communications. 2022;13(1). doi:10.1038/s41467-022-32402-0.

32. Silué DK, Díaz de León Derby M, Delahunt CB, Le Ny AL, Spencer E, Armstrong M, et al. A schistosomiasis dataset with bright- and darkfield images. MICCAI. 2024;.

33. WHO Expert Committee on the Control of Schistosomiasis. Prevention and Control of Schistoso-miasis and Soil-Transmitted Helminthiasis: Report of a WHO Expert Committee;. https://iris.who.int/handle/10665/42588.

34. Pedregosa F, Varoquaux G, Gramfort A, Michel V, Thirion B, Grisel O, et al. Scikit-learn: Machine Learning in Python. Journal of Machine Learning Research. 2011;12:2825–2830.

35. Delahunt CB, Gachuhi N, Horning MP. Metrics to guide development of machine learning algorithms for malaria diagnosis. Frontiers in Malaria. 2024;2. doi:10.3389/fmala.2024.1250220.

36. Subbaswamy A, Saria S. From development to deployment: dataset shift, causality, and shift-stable models in health AI. Biostatistics. 2019;21(2):345–352. doi:10.1093/biostatistics/kxz041.

37. Balagopalan A, Baldini I, Celi LA, Gichoya J, McCoy LG, Naumann T, et al. Machine learning for healthcare that matters: Reorienting from technical novelty to equitable impact. PLOS Digital Health. 2024;3(4):e0000474. doi:10.1371/journal.pdig.0000474.

38. Das D, Vongpromek R, Assawariyathipat T, Srinamon K, Kennon K, Stepniewska K, et al. Field evaluation of the diagnostic performance of EasyScan GO: a digital malaria microscopy device based on machine-learning. Malaria Journal. 2022;21(1). doi:10.1186/s12936-022-04146-1.

39. Moya Muñoz GG, Brix O, Klocke P, Harris PD, Luna Piedra JR, Wendler ND, et al. Single-molecule detection and super-resolution imaging with a portable and adaptable 3D-printed microscopy platform (Brick-MIC). bioRxiv. 2024; doi:10.1101/2023.12.29.573596.

