## Supplementary figures and images for "Multi-contrast machine learning improves schistosomiasis diagnostic performance"

### Supplementary Figure 1

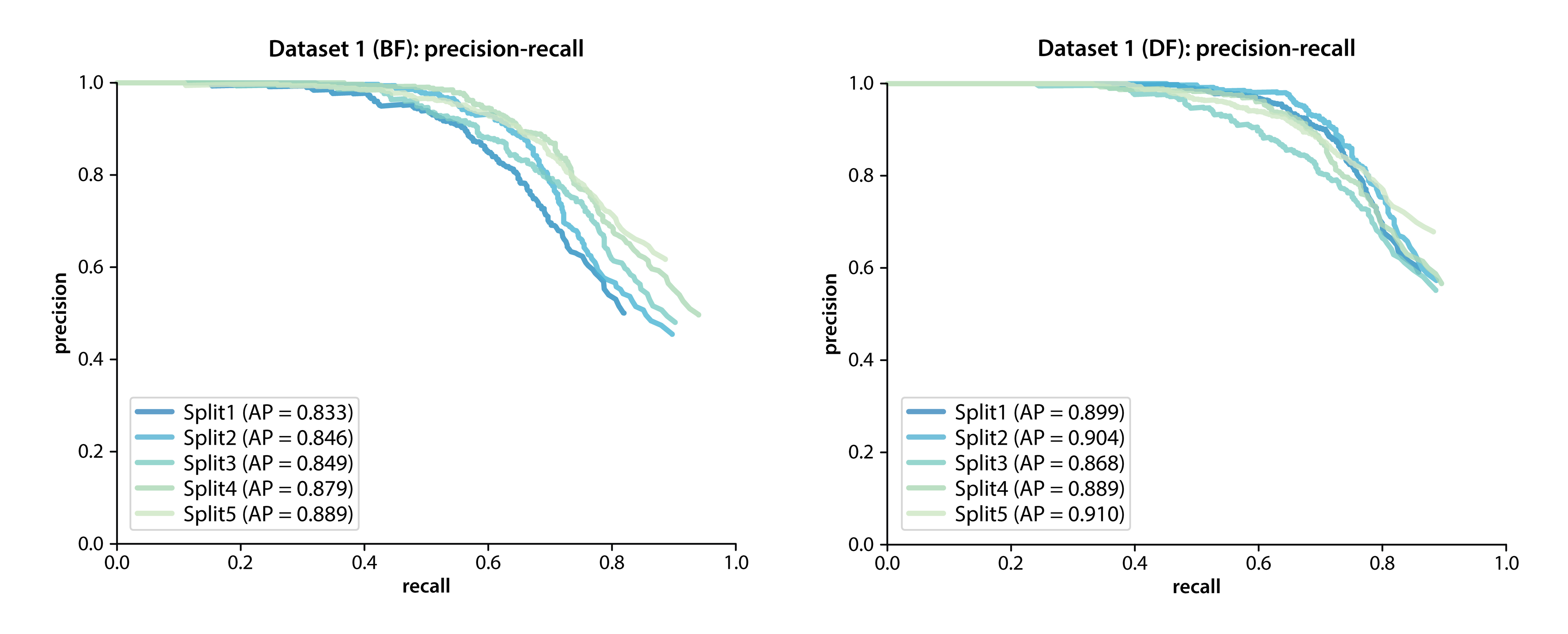

### Supplementary Figure 2

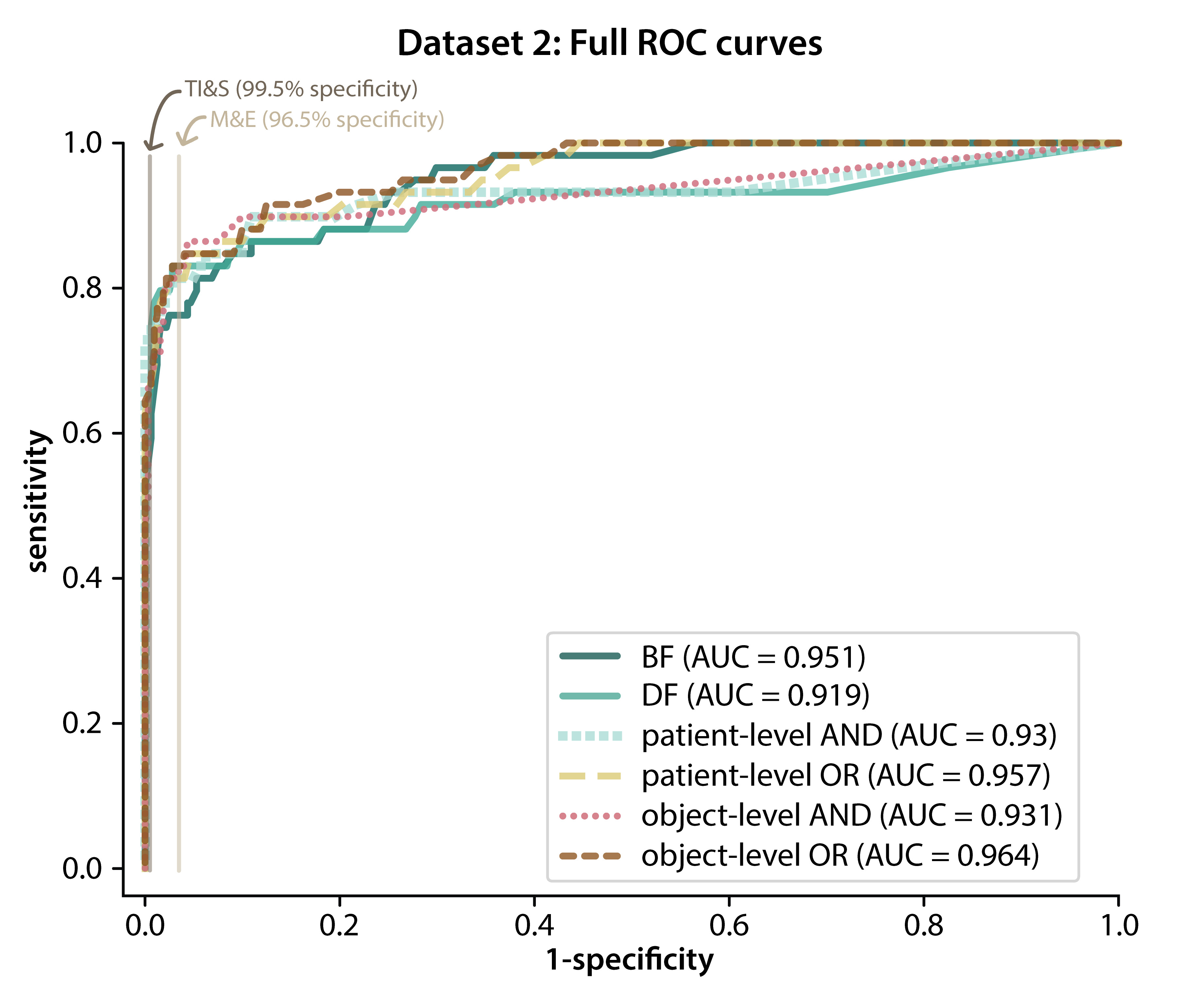
